# Unsuspected consequences of synonymous and missense variants in *OCA2* can be detected in blood cell RNA samples of patients with albinism

**DOI:** 10.1101/2023.07.07.23292371

**Authors:** Vincent Michaud, Angèle Sequeira, Elina Mercier, Eulalie Lasseaux, Claudio Plaisant, Smail Hadj-Rabia, Sandra Whalen, Dominique Bonneau, Anne Dieux-Coeslier, Fanny Morice-Picard, Juliette Coursimault, Benoît Arveiler, Sophie Javerzat

**Author notes:** both authors contributed equally to this work and should be considered first authors (Vincent Michaud, Angèle Sequeira).

## Abstract

Oculocutaneous albinism type 2 (OCA2) is the second most frequent form of albinism and represents about 30% of OCA worldwide. As with all types of OCA, patients present with hypopigmentation of hair and skin as well as severe visual abnormalities. We focused on a subgroup of 29 patients for whom genetic diagnosis was pending because at least one of their identified variants in or around exon 10 of *OCA2* is of uncertain significance (VUS). By minigene assay, we investigated the effect of these VUS on exon 10 skipping and showed that not only intronic but also some synonymous variants can result in enhanced exon skipping. We further found that excessive skipping of exon 10 could be detected directly on blood samples of patients and of their one parent with the causal variant, avoiding invasive skin biopsies. Moreover, we show that variants which result in lack of detectable *OCA2* mRNA can be identified from blood samples as well, as shown for the most common *OCA2* pathogenic missense variant c.1327G>A/p.(Val443Ile). In conclusion, blood cell RNA analysis allows testing the potential effect of any *OCA2* VUS on transcription products. This should help to elucidate yet unsolved OCA2 patients and improve genetic counseling.

**SIGNIFICANCE:** Our study allows us to reconsider variants of unknown significance of *OCA2* as pathogenic as they induce exon 10 skipping. We show that mis-splicing as well as other types of transcripts imbalance can be detected directly from blood cell samples, avoiding invasive biopsies. We recommend systematic collection of a blood RNA sample from patients with inconclusive genetic diagnosis and suspected OCA2 (e.g., only one pathogenic variant in *OCA2*; 1 VUS; 2 VUS in *trans*).

## INTRODUCTION

Molecular diagnosis of patients with clinical suspicion of albinism is essential to confirm the type of albinism, adapt care of patients and offer genetic counseling to the families. Twenty-one genes are currently known to cause albinism, yet ∼30% of patients remain genetically unsolved with a significant proportion of them (60%) presenting at least one variant of uncertain significance (VUS) in one of the known albinism genes (Lasseaux et al., 2018).

Truncating variants are usually easy to classify depending on the site of deletion in the protein. For missense variants, classification is mainly based on frequency and *in-silico* prediction tools. Functional assays are required when the prediction is of low confidence. For rare synonymous variants, a pathogenic effect on exon splicing regulation or codon usage can be sought *in-silico*, but prediction tools are poorly efficient for such interferences (Lord & Baralle, 2021).

Oculocutaneous albinism type 2 (OCA2) is the second most frequent form of albinism and represents about 30% of OCA worldwide (Kamaraj & Purohit, 2014). Like all other OCAs, it is autosomal recessive and characterized by general hypopigmentation of the hair, skin and eye, as well as visual deficiencies. OCA2 results from pathogenic variants in the *OCA2* gene, formerly known as *P* gene.

Our albinism patient cohort (n=1900) includes 574 individuals with at least one rare variant detected in the coding sequence of *OCA2* (30%), reflecting the reported worldwide frequency of OCA2 albinism (Jaworek et al., 2012). As expected from the important size of the gene (345 kb), a substantial amount of rare variants are not robustly classified as pathogenic and consequently, many suspected OCA2 patients present with at least one VUS.

In Human, *OCA2* is located on chromosome 15q11-q13 and has 24 exons. The encoded P protein comprises 838 amino acids. It localizes at the lysosomal and lysosomal-like (melanosomal) membranes through its 12 transmembrane domains (Bellono et al., 2014). The P protein plays an essential role in chloride conductance that results in optimal melanosomal pH ensuring correct pigment production and assembly in melanocytes as well as in the retinal pigment epithelium (Bellono et al., 2014). Additionally, loss of function of *OCA2* has been reported to be associated with disruption of the Unfolded Protein Response (Cheng et al., 2013).

At the level of mRNAs, *OCA2* transcripts are found as two major forms: the full-length transcript (3143 nt; MANE transcript, Refseq NM_000275, Ensembl <u>ENST00000354638.3</u>, CCDS10020), here referred to as Ex10^+^, encoding the P protein, and an alternative coding transcript deleted of the in-frame exon 10 (72 bp encoding 24 amino acids, 3071 nt; Refseq NM_001300984, Ensembl <u>ENST00000353809.5</u>), here referred to as Ex10^del^. In most healthy tissues that express significant levels of *OCA2* mRNA (skin, retina, brain, arteries), the alternative form Ex10^del^ is detected in lesser quantities than the functional Ex10^+^ (Jaworek et al., 2012; GTEx Portal: https://gtexportal.org/). Of note, the Ex10^del^ putative protein has not been described but is predicted non-functional as it would lack its entire 3^rd^ transmembrane domain resulting in inversion of intra to extra-melanosomal membrane of the N-terminal end of the protein (http://wlab.ethz.ch/protter/). In 2012, Jaworek et al. identified a pathogenic variant, c.1045-15T>G (intron 9), homozygous in five Pakistani families presenting with oculocutaneous albinism. This intronic single nucleotide polymorphism (SNP) was shown to induce full skipping of exon 10 in a minigene assay (Jaworek et al., 2012).

From our albinism patient cohort, we selected 29 patients bearing variants lying in or around exon 10. These variants, classified either pathogenic or of uncertain significance, were investigated for their effect on splicing. By minigene assay, we find that not only intronic SNPs but also synonymous variants and an in-frame deletion in exon 10 can result in enhanced exon skipping to an extent that supports pathogenicity. We also show that both transcripts of *OCA2* can be detected in RNAs extracted from control blood cell samples with proportions equivalent to what is described in skin. This enables us to test excessive exon 10 skipping directly from blood samples rather than from more invasive skin biopsies.

Finally, we find that not only exon skipping but also *OCA2* mRNA levels can be assessed from blood samples. Indeed, we show that two unrelated patients carrying the *OCA2* pathogenic variant c.1327G>A, predicted as missense p.(Val443Ile) had no corresponding *OCA2* mRNA detected in blood samples. *OCA2* mRNA analysis from blood RNA samples can therefore provide valuable information on variants resulting in transcript reduction or instability that may lie in deep intronic regions and are left undetected by conventional exon and intron/exon junction sequencing.

## MATERIALS AND METHODS

### Editorial Policies and Ethical Considerations

This study was approved by our local ethics committee: Comité de Protection des Personnes Bordeaux Outre-Mer III. Written informed consent was received from the patients and the participating families according to the declaration of Helsinki principles of medical research involving human subjects.

### Patients

Patients were referred for a clinical suspicion of albinism by ophthalmologist, dermatologist, hematologist or geneticist for a molecular analysis. Sequencing was performed by the Department of Medical Genetics, University Hospital of Bordeaux using Next Generation Sequencing of all known albinism genes.

### Variant predictions

Variant predictions were gathered from MobiDetails (Baux et al., 2021); SpliceAI (Jaganathan et al., 2019); SPiP (Leman et al., 2022), and CADD (Kircher et al., 2014).

### Data availability statement

Novel variants have been uploaded to ClinVar public database.

### Minigene constructs and splicing assay

Genomic DNA was extracted from peripheral blood leukocytes of the heterozygous mother (M1) from Family 1 carrying the c.1080C>T variant. Both the wild-type and the c.1080C>T sequence of exon 10 flanked by 327bp of intron 9 and 429bp of intron 10 were PCR amplified and cloned in the minigene assay vector pSPL3B by homologous recombination (Burn et al., 1995). PCR products were prepared using the “In-Fusion HD Cloning kit with Cloning Enhancer Treatment” (Takara Bio) and recombined with Not I linearized pSPL3B vector. Variants of interest were introduced into the wild-type cloned sequence with QuikChange Site-Directed Mutagenesis Kit (Agilent) following the manufacturer’s recommendations. Mutagenesis primers were designed as recommended. All constructs were confirmed through direct sequencing (Eurofins) followed by alignment with the reference *OCA2* sequence (ENSG00000104044, www.ensembl.org). All primers are listed in table S1.

HeLa cells were cultured in DMEM +GlutaMAX^TM^ (Gibco) supplemented with 1% penicillin and streptomycin and 10% fetal bovine serum. Transfections of HeLa cells were performed in 6-well plates using FuGENE® HD Transfection Reagent (Promega) with 3 µg of DNA and 1:1 ratio according to the manufacturer’s recommendations. Forty-eight hours after transfection, total RNAs were purified using the NucleoSpin RNA Mini kit (Macherey Nagel). First-strand cDNA synthesis was carried out through Oligo(T)-primed reverse transcription using M-MLV reverse transcriptase (Promega). The resulting cDNA was amplified by PCR using SD6 and SA2 vector-specific primers (listed in table S1). The PCR amplification reaction was performed in 20 μL using the Q5 High-fidelity Master kit (New England Biolabs). PCR conditions were 35 cycles (95°C for 15 s, 60°C for 15 s, and 72°C for 20 s), allowing amplification to remain in the exponential range. PCR products were separated by electrophoresis through 1.5% agarose gel, checked by DNA sequencing (Eurofins) and each band signal was quantified by the ImageJ software (https://imagej.nih. gov/ij/, RRID: nif-0000-30467). Quantification of the abnormal splicing percentage was densitometrically calculated as the percentage of exclusion (%) = (lower band/[lower band + upper band]) × 100. Error bars represent SEM (*n* = 3). ∗ indicates *p* < 0.05, unpaired Student’s *t*-test.

### Allele-specific cloning analysis from biopsies

Skin biopsies from Family 1 (patient and healthy parents) were disrupted and processed for total RNA extraction using Rneasy Minikit (QIAGEN) including on-column DNase treatment following manufacturer’s instructions. 350ng-600ng of total RNA were retro-transcribed as described above. Based on the reference sequence NM_000275.3 (http://www.ensembl.org), a cDNA fragment was PCR-amplified using primers located in exons 7-8 boundary and in exon 16 of the gene (Table S1). The resulting amplicon includes a SNP (dbSNP, rs1800411) present in exon 15 allowing to discriminate maternal or paternal inheritance. Amplicons were cloned in pMiniT vector using the PCR Cloning Kit (New England Biolabs) and sequenced.

### RT-PCR on blood samples

Blood samples were collected in PAXgene® tubes and directly processed within the next 4 days or later after storage at −20°C. Total RNA extraction was performed using the PAXgene® Blood RNA kit (QIAGEN). Two µg of total RNA were retro-transcribed using M-MLV reverse transcriptase (Promega) and specific expression of *OCA2* was assessed by PCR amplification (primers listed in table S1). Touch-down PCR was performed for 10 cycles with Tm ranging from 65 to 55°C and followed by 35 cycles 95°C for 15 s, 62°C for 15 s, and 72°C for 30 s and the resulting fragments (383 bp with and 311 bp without exon 10) were quantified after electrophoresis. For the c.1327G>A study, the RT-PCR product containing exon 13 (298 bp) was sequenced (Eurofins) or submitted to MaeIII digestion that discriminates wild-type (1 cut: 126bp+172bp) from variant (no cut: 298 bp) sequence at position c.1327 (Lee et al, 1994). MaeIII digestion was carried out for 1h at 37°C after purification of the amplicons using the PCR Clean Up kit (Qiagen).

## RESULTS

From our cohort, we identified 414 individuals with 2 rare variants in *OCA2*. Here we selected 29 patients, with at least one rare variant lying in or around exon 10. Ninety percent of the selected patients with available clinical data (18/20) presented with hypopigmentation of the hair and skin (Table 1). Ocular features of albinism were frequent to systematic: transillumination in 91% of patients with available clinical data (10/11), retinal hypopigmentation in 100% (14/14), foveal hypoplasia in 67% (4/6) and nystagmus in 92% (22/24). Visual evoked potentials were not available for most patients.

**Table 1:** Genotype and phenotype of patients included in this study. Nomenclature based on NM_000275.3 HTZ: heterozygous; HMZ: homozygous; ND: Not determined; NA: Not applicable; Y: Yes ^#^already published in Lasseaux *et al* 2018 ^§^already published in Rooryck *et al* 2008 Patients for which RNA analysis has been performed in this study are highlighted in grey

| PATIENT | VARIANTS OCA2 EXON 10 |  |  | SECOND OCA2 VARIANT |  |  | PARENTAL<br>SEGREGATION | SEX | ORIGIN | SKIN COLOR | HAIR COLOR | IRIS COLOR | IRIS<br>TRANSLUMINATION | RETINAL<br>HYPOPIGMENTATION | FOVEAL<br>HYPOPLASIA | NYSTAGMUS | PHOTOPHOBIA | LOW VISUAL ACUITY |
| --- | --- | --- | --- | --- | --- | --- | --- | --- | --- | --- | --- | --- | --- | --- | --- | --- | --- | --- |
|  | c. | p. | Status | c. | p. | Status |  |  |  |  |  |  |  |  |  |  |  |  |
| P1 | 1045-10T>G | NA | HTZ | 2330G>A | Cys777Tyr | HTZ | Y | F | France | white | platine white | blue | + | + | ND | + | ND | + |
| P2<br>(SIBLING OF P1) | 1045-10T>G | NA | HTZ | 2330G>A | Cys777Tyr | HTZ | Y | F | France | white | platine white | ND | ND | ND | ND | + | ND | ND |
| P3<sup>#</sup> | 1045-9T>G | NA | HOM | - | - | - | Y | M | New Caledonia | ND | platine white | ND | ND | ND | ND | + | ND | ND |
| P4 | 1045-9T>G | NA | HTZ | 1064C>A | Ala355Glu | HTZ | Y | M | New Caledonia | ND | platine white | ND | ND | ND | ND | + | ND | ND |
| P5 | 1045-9T>G | NA | HTZ | Deletion exon 7 | - | HTZ | Y | M | ND | cream | Blond | blue | ND | ND | ND | + | + | ND |
| P6 | 1047C>T | Ile349= | HTZ | 1874A>G | Lys625Arg | HTZ | Y | M | France | pink | Pale brown | brown | + | + | ND | + | - | ND |
| P7<sup>#</sup> | 1064C>A | Ala355Glu | HTZ | 2324G>A | Gly775Asp | HTZ | Y | F | New Caledonia | ND | ND | ND | ND | ND | ND | ND | ND | ND |
| P8<sup>#</sup> | 1064C>A | Ala355Glu | HTZ | 2324G>A | Gly775Asp | HTZ | ND | M | New Caledonia | ND | ND | ND | ND | ND | ND | ND | ND | ND |
| P9 | 1064C>A | Ala355Glu | HTZ | 2324G>A | Gly775Asp | HTZ | ND | M | New Caledonia | ND | ND | ND | ND | ND | ND | + | ND | ND |
| P10<sup>#</sup> | 1076G>A | Gly359Asp | HTZ | 1983G>T | Leu661Phe | HTZ | ND | F | Algeria | ND | ND | ND | ND | ND | ND | ND | ND | ND |
| P11 | 1076G>A | Gly359Asp | HOM | - | - | - | Y | F | Algeria | white | yellow | blue | ND | ND | ND | + | + | ND |
| P12<sup>#</sup><br>Family 2 | 1076G>A | Gly359Asp | HTZ | 2359G>A | Ala787Thr | HTZ | Y | M | Algeria | white | platine white | blue | ND | + | ND | + | + | + |
| P13 | 1076G>A | Gly359Asp | HTZ | 2359G>A | Ala787Thr | HTZ | ND | F | Algeria | white | platine white | blue | + | + | + | + | + | + |
| P14 | 1080C>T | Ser360= | HTZ | 516-3C>A | ? | HTZ | ND | M | France | ND | ND | ND | ND | ND | ND | ND | ND | ND |
| P15 | 1080C>T | Ser360= | HTZ | 2330G>A | Cys777Tyr | HTZ | Y | F | France | white | platine white | blue | + | + | ND | + | + | ND |
| **P16** | 1080C>T | Ser360= | HTZ | 1349C>T | Thr450Met | HTZ | Y | F | France | white | yellow | blue | + | + | ND | + | ND | + |
| **P17** | 1080C>T | Ser360= | HTZ | None | - | - | ND | M | France | ND | ND | ND | ND | ND | ND | + | ND | + |
| **P18**<br>**Family 3** | 1080C>T | Ser360= | HTZ | 1327G>A | Val443Ile | HTZ | Y | F | France | cream | yellow | blue | + | + | - | + | + | + |
| **P19**<br>**(SIBLING OF**<br>**P18)**<br>**Family 3** | 1080C>T | Ser360= | HTZ | 1327G>A | Val443Ile | HTZ | Y | M | France | cream | yellow | green | + | + | - | + | + | + |
| **P20** | 1080C>T | Ser360= | HTZ | None | - | - | ND | F | France | pink | red | ND | ND | + | + | - | - | + |
| **P21** | 1080C>T | Ser360= | HTZ | 1778T>C | Phe593Ser | HTZ | Y | M | France | ND | ND | ND | ND | ND | ND | + | + | - |
| **P22**<br>**Family 1** | 1080C>T | Ser360= | HTZ | 2433G>T | Arg811Ser | HTZ | Y | F | France | white | yellow | blue | + | + | + | + | + | + |
| **P23<sup>#</sup>** | 1095_1103del | Ala366_368del | HTZ | 1481C>T | Ser494Phe | HTZ | Y | M | Marocco | cream | yellow | blue | + | + | ND | + | + | ND |
| **P24** | 1095_1103del | Ala366_368del | HTZ | 2339G>A | Gly780Asp | HTZ | ND | M | Mali | cream | yellow | ND | ND | ND | ND | + | ND | ND |
| **P25** | 1103C>T | Ala368Val | HTZ | Deletion<br>exon 7 | - | HTZ | Y | M | Cameroun | ND | ND | ND | ND | ND | ND | ND | + | + |
| **P26** | 1103C>T | Ala368Val | HTZ | Deletion<br>exon 7 | - | HTZ | ND | M | Gabon | white | light<br>brown | brown | - | + | + | - | + | ND |
| **P27** | 1103C>T | Ala368Val | HTZ | 2089C>T | His697Tyr | HTZ | Y | F | France | ND | ND | brown | ND | ND | ND | + | ND | ND |
| **P28** | 1103C>T | Ala368Val | HOM | - | - | - | ND | F | Antilles | cream | yellow | Grey | ND | + | ND | + | + | ND |
| **P29<sup>#,§</sup>** | 1116+6T>C | NA | HTZ | 1327G>A | Val443Ile | HTZ | ND | F | France | white | yellow | blue | + | + | ND | + | + | + |

In 27 out of 29 patients, two rare variants were detected in the coding sequence of *OCA2*: 3 patients were homozygotes (P3, P11, P28), 24 had two different variants out of which 16 patients could be confirmed compound heterozygotes by segregation analysis. Two patients (P17 and P20) had only one heterozygous variant in *OCA2* after coding sequence analysis. Interestingly, the same compound heterozygous genotypes were detected in unrelated patients of the same geographic origin: c.1064C>A/p.(Ala355Glu); c.2324G>A/p.(Gly775Asp) from New Caledonia (in P7, P8, P9) and c.1076G>A/p.(Gly359Asp); c.2359G>A/p.(Ala787Thr) from Algeria (P14, P15). Of note, the most frequent *OCA2* pathogenic variant c.1327G>A/p.(Val443Ile) (f=0.003 in gnomAD) was present in 2 siblings (P18, P19) of the cohort as well as one unrelated patient (P29).

In total, the 29 selected patients carry 9 different rare variants in or around exon 10 of *OCA2* (3 intronic, 3 missense, 2 synonymous and 1 in-frame deletion), that display very low frequency in population databases. As detailed in Table 2, three were absent from both versions of gnomAD databases, 4 were very rare with less than 10 heterozygotes, 1 was found in 37 heterozygotes but no homozygotes. Only c.1103C>T/p.(Ala368Val) was found as homozygous in gnomAD (1/210 000 genomes). These frequencies are compatible with pathogenicity in a rare autosomal recessive disease. Of note, high probability of mis-splicing was not predicted by *in silico* tools for any of the variants.

**Table 2:** Characteristics of OCA2 exon 10 variants. Nomenclature based on NM_000275.3; HTZ: heterozygous; HMZ: homozygous; NA: Not applicable Predictions and frequency gathered from MobiDetails https://mobidetails.iurc.montp.inserm.fr/MD/; Splice AI : https://spliceailookup.broadinstitute.org/; SPiP : https://github.com/raphaelleman/SPiP; CADD : https://cadd.gs.washington.edu/snv; gnomAD : https://gnomad.broadinstitute.org/; Clinvar : https://www.ncbi.nlm.nih.gov/clinvar/.

| OCA2 Variants |  | In Silico Predictions |  |  | gnomAD allele frequency<br>(Number HTZ and HMZ) |  | Clinvar |
| --- | --- | --- | --- | --- | --- | --- | --- |
| c. | p. | SpliceAI | SPiP | CADD<br>phred | v2 | v3 |  |
| 1045-10T>G | NA | 0,19<br>(below threshold) | 23%<br>(low alteration ) | 14,85 | 0 | 0 | Absent |
| 1045-9T>G | NA | 0,24<br>(low effect) | 23%<br>(low alteration ) | 8,14 | 0.00002126<br>(6 HTZ) | 0.000006570<br>(1 HTZ) | Uncertain significance |
| 1047C>T | Ile349= | 0,04<br>(below threshold) | 34%<br>(low ESE alteration) | 0,08 | 0.00008146<br>(23 HTZ) | 0.00009200<br>(14 HTZ) | Likely benign |
| 1064C>A | Ala355Glu | 0,00<br>(no effect) | 35%<br>(complex alteration) | 25,1 | 0 | 0 | Absent |
| 1076G>A | Gly359Asp | 0,02<br>(below threshold) | 3%<br>(no effect) | 24 | 0.000007964<br>(2 HTZ) | 0 | Pathogenic |
| 1080C>T | Ser360= | 0,02<br>(below threshold) | 23%<br>(low site creation) | 3,66 | 0.00001770<br>(5 HTZ) | 0.00001314<br>(2 HTZ) | Uncertain significance<br>Likely benign |
| 1095_1103del | Ala366_368del | 0,08<br>(below threshold) | 8%<br>(no effect) | 17 | 0 | 0.000006568<br>(1 HTZ) | Absent |
| 1103C>T | Ala368Val | 0,01<br>(below threshold) | 47%<br>(low ESE alteration) | 25,4 | 0.0002584<br>(73 HTZ) | 0.0007687<br>(115 HTZ ; 1 HMZ) | Pathogenic<br>Likely Pathogenic<br>Uncertain significance |
| 1116+6T>C | NA | 0,41<br>(low effect) | 98%<br>(high alteration) | 19,05 | 0 | 0 | Absent |

### Both intronic and exonic variants can result in *OCA2* exon 10 skipping as shown by minigene assay

The variants described in Table 2 were tested in a minigene splicing assay as illustrated in Figure 1A. *OCA2* exon 10 (*OCA2*-Ex10) (72 bp) flanked by ∼400 bp of intronic sequences was cloned from a healthy heterozygote parent genomic DNA in pSPL3B. Transfection of wild-type minigene (pSPL3B-Ex10-WT) in HeLa cells followed by RT-PCR amplification generated fragments of the expected size: a major band at 335 bp corresponding to correct splicing and a minor band (around 5%) at 263 bp corresponding to skipping of *OCA2*-Ex10 reflecting the splicing pattern of *OCA2* in most human tissues (GTEx databases). The intronic variant c.1045-15T>G described as pathogenic in homozygous patients by Jaworek et al. (2012) was included as a reference and, as expected, generated only fragments corresponding to skipping of *OCA2*-Ex10. The three intronic VUS that were tested, whether in intron 9 (c.1045-10T>G, c.1045-9T>G) or in intron 10 (c.1116+6T>C), equally resulted in systematic skipping of *OCA2*-Ex10 and can therefore be classified pathogenic. None of the three missense variants of *OCA2*-Ex10 (c.1064C>A/p.(Ala355Glu), c.1076G>A/p.(Gly359Asp), c.1103C>T/p.(Ala368Val)) generated excess of skipped products which indicates that their pathogenicity is likely due to the amino acid change in the translated protein as predicted *in-silico* (Table 2). In contrast, both synonymous variants (c.1047C>T/p.(Ile349=) and c.1080C>T/p.(Ser360=)) enhanced skipping of *OCA2*-Ex10 that was confirmed by sequencing of cloned RT-PCR products of both sizes (335 and 263 bp). Increased skipping was also obtained for a 9 bp in-frame deletion (c.1095_1103del/p.(Ala366_368del)) (Figure 1B). Variant c.1080C>T/p.(Ser360=) was found in 8 unrelated patients of our cohort (Table 1), 6 of whom were compound heterozygotes with a pathogenic variant *in trans* (P14, P15, P16, P18 and sibling P19, P21, P22) and 2 with no second identified *OCA2* variant (P17, P20). Of those, P22 (c.1080C>T; c.2433G>T) (Family 1) was further studied as cutaneous biopsies were available for this patient, as well as for her mother (M1) who carries the c.1080C>T/p.(Ser360=) variant and for her father (F1) who carries the pathogenic missense variant c.2433G>T/p.(Arg811Ser) (Figure 1C). Sequencing of cloned RT-PCR products from each of the 3 biopsies indicated a higher proportion of skipped Ex10^del^ transcripts in P22 and her mother compared to her father for whom the two RT-PCR products with or without exon 10 were cloned in equivalent quantities. Moreover, the 5/26 independent clones (19.2%) corresponding to normal splicing that could be recovered from patient P22 biopsy were all of paternal origin (c.1080C which is in *cis* of c.2433G>T/p.(Arg811Ser)) indicating major reduction or absence of functional *OCA2* transcripts in this patient.

**Figure 1:**
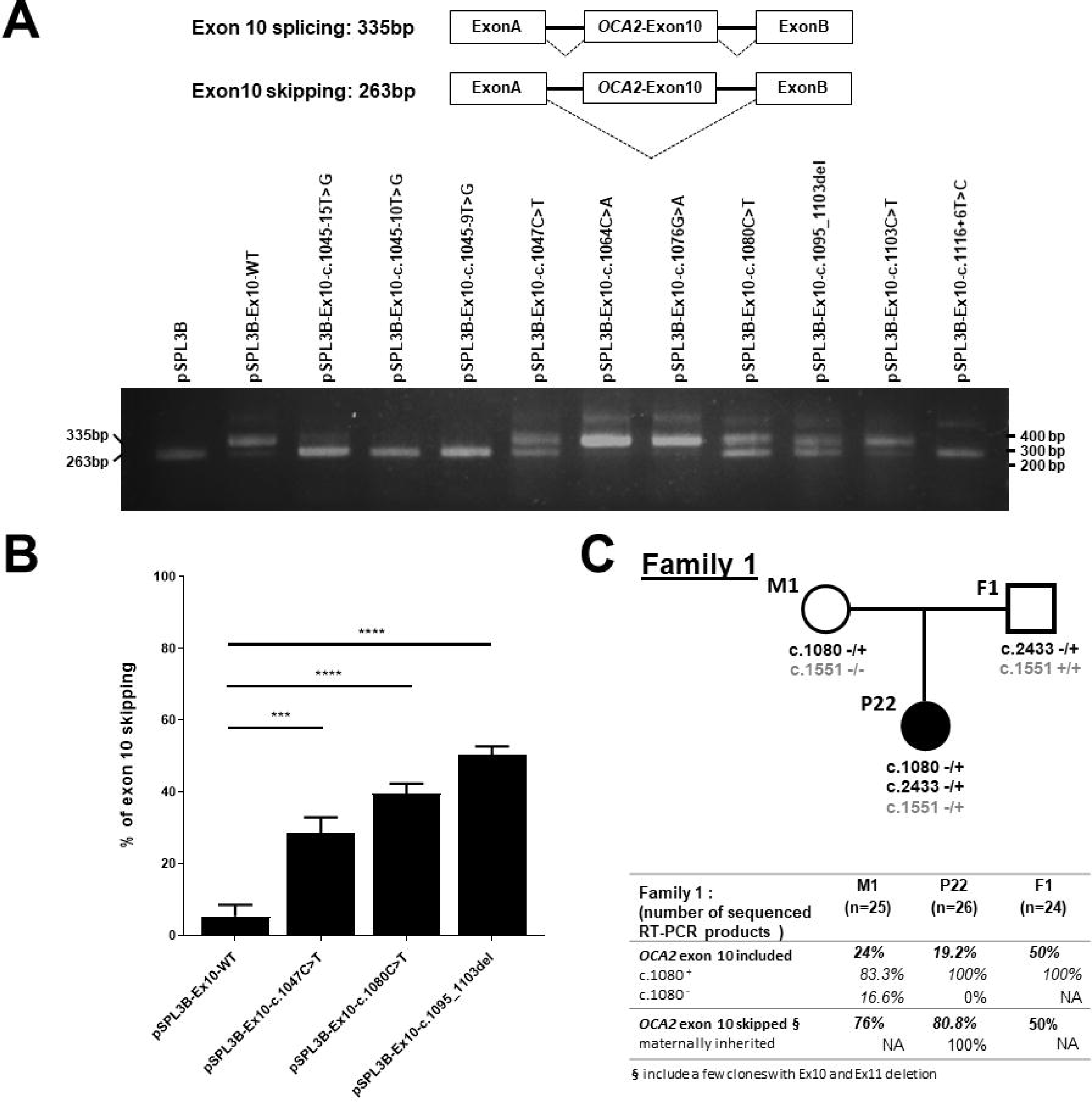
Functional analysis of selected variants in or around *OCA2* exon 10. **A**: Representative scheme of the minigene assay with two expected PCR product sizes with correct splicing or skipping of exon 10 (upper panel). Agarose gel electrophoresis of RT-PCR products from minigene assay of 10 variants compared to wild-type (lower panel). **B**: Relative quantification of exon 10 skipping induced by 2 different synonymous variants and one in-frame deletion compared to WT showing significantly increased skipping. Error bars represent SEM (*n* = 3). ∗ indicates *p* < 0.05, unpaired Student’s *t*-test. **C**: For Family 1, skin biopsies were obtained from patient P22, her mother M1 and her father F1. RT-PCR products encompassing the exon 10 region were cloned and sequenced to determine exon inclusion or exclusion. Among clones with exon 10, those bearing the reference allele c.1080C (c.1080^+^) or bearing variant c.1080C>T (c.1080^−^) were scored. Father F1 does not carry variant c.1080C>T so that no c.1080^−^ RT-PCR products are expected (non applicable, NA). For patient P22, all clones with exon 10 included had the c.1080^+^ paternal sequence. To determine the origin of clones with skipped exon 10, the SNP c.1551C>T (rs1800411, genotypes indicated in grey) was sequenced which indicated that they were all expressed from the maternal allele with variant c.1080C>T (c.1080^−^).

In total, the minigene assay run on 9 selected variants in or around *OCA2* exon 10, functionally supports pathogenicity due to enhanced exon skipping for all of the 6 variants that are not missense: 3 intronic variants (c.1045-10T>G; c.1045-9T>G; c.1116+6T>C), two synonymous variants (c.1047C>T/p.(Ile349=); c.1080C>T/p.(Ser360=)) and a 9 bp in-frame deletion c.1096_1103del/p.(Ala366_368del), all 6 being absent or classified VUS in ClinVar.

### *OCA2* VUS can be assayed for their impact on functional mRNA production from blood cell samples

Unlike the major OCA gene *TYR* encoding tyrosinase, *OCA2* expression is not restricted to pigment cells. It is supposed to be slightly expressed in blood cells according to the GTEx expression database (https://www.gtexportal.org/home/gene/OCA2). Focusing on *OCA2*-Ex10 sensitivity to sequence modification resulting in exon skipping, we therefore asked if enhanced *OCA2*-Ex10 skipping could be detected from *OCA2* transcripts extracted from patients’ blood samples rather than from more invasive skin biopsies. Blood samples collected in PAXgene® tubes allowing recovery of mRNAs were available for two families (Family 2: mother M2 and father F2 of patient P12 and Family 3: mother M3 and father F3 of patient siblings P18 and P19) as well as for one unrelated patient (P29). These blood RNA samples were used to test *OCA2* mRNA detection and to further assess *OCA2*-Ex10 skipping detection.

The first case studied is patient P12 (Family 2) who inherited the pathogenic *OCA2*-Ex10 missense variant c.1076G>A/p.(Gly359Asp) from his father (F2) and a second pathogenic missense variant c.2359G>A/p.(Ala787Thr) in exon 23 from his mother (M2) (Figure 2A). The c.1076G>A variant is not expected to modify the balance of exon 10 skipping based on minigene assay results (Figure 1A). Firstly, we successfully amplified specific *OCA2* amplicons using one primer in exon 9 and the second primer in exon 12 (Table S1), confirming that RT-PCR from blood RNA samples is reliable. Secondly, both products corresponding to *OCA2* Ex10^+^ and Ex10^del^ were detected. Considering M2 as a control for normal balance of the 2 transcripts extracted from blood cells (with equivalent quantities of both fragments detected), we show that the Ex10^del^ fragment is not over-represented in P12 and F2 carrying the c.1076G>A variant. This observation confirms the minigene assay results indicating that missense variant c.1076G>A has no effect on splicing *in vivo* in blood cells.

**Figure 2:**
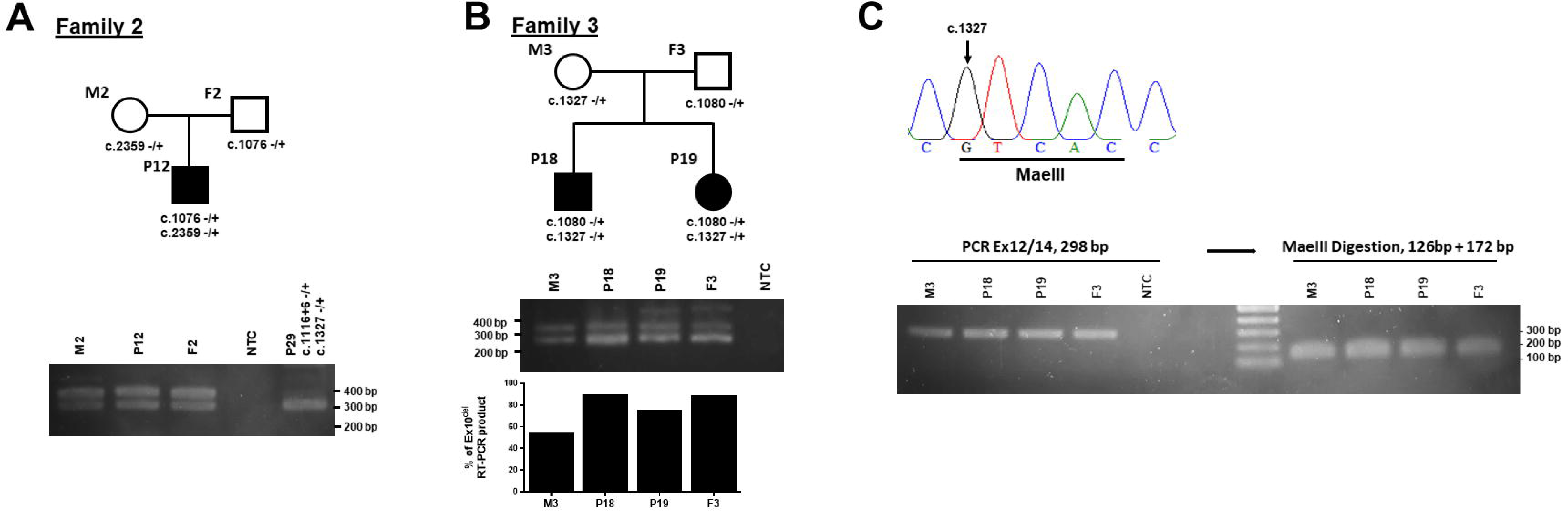
OCA2 exon 10 skipping detection and consequence of a missense variant on mRNA from blood cells. **A**: Pedigree of Family 2 showing segregation of 2 pathogenic missense variants found in patient P12 (upper panel). RT-PCR products of *OCA2* exon 10 from blood cells RNAs (primers in exon 9 and 12, see Table S1). From Family 2, normal balance of the two types of transcripts *OCA2*-Ex10^+^ and *OCA2*-Ex10^del^ in both parents and affected child with no over-representation of *OCA2*-Ex10^del^ as expected. By contrast, only *OCA2*-Ex10^del^ derived RT-PCR products are detected in patient P29 that carries the intronic c.1116+6T>C variant (lower panel). NTC: Non-template control **B**: Pedigree of Family 3 showing the inheritance of the variants found in patients P18 and P19, c.1327G>A from mother M3, c.1080C>T from father F3 (upper panel). RT-PCR products of *OCA2 e*xon 10 from blood cells RNAs for Family 3 members showing normal balance of *OCA2*-Ex10^+^ vs. *OCA2*-Ex10^del^ RT-PCR products for M3 whereas *OCA2*-Ex10^del^ is over-represented in samples from P18, P19 and F3 all bearing *OCA2* the c.1080C>T synonymous variant (middle panel). Relative quantification of *OCA2*-Ex10^del^ RT-PCR products (lower panel). **C**: Electrophoregram showing MaeIII enzyme restriction site localisation that is removed by *OCA2* exon 13 variant c.1327G>A (upper panel). RT-PCR products of OCA2 exon 13 primers in exon 12 and 14, see Table S1) before and after digestion by MaeIII restriction enzyme for 4 members of Family 3 showing complete digestion of PCR products for F3 bearing WT c.1327G but also for P18, P19 and F3 bearing c.1327G>A variant (lower panel).

We next asked if blood samples might directly be used for investigating enhanced splicing of *OCA2*-Ex10. Messenger RNAs were recovered from a blood sample of P29 who is compound heterozygous for the intronic variant c.1116+6T>C in *trans* with the most frequent *OCA2* pathogenic variant c.1327G>A/p.(Val443Ile) located in exon 13 (*OCA2*-Ex13). An over-representation of *OCA2*-Ex10^del^ was expected due to variant c.1116+6T>C which was shown to induce complete exon 10 skipping by minigene assay whereas c.1327G>A/p.(Val443Ile) should in theory allow both *OCA2*-Ex10 and *OCA2*-Ex10^del^ to be generated. Unexpectedly, only the *OCA2*-Ex10^del^ amplicon was detected from this patient (Figure 2A) as confirmed by sequencing (not shown), suggesting a significant imbalance of the transcripts derived from the c.1327G>A carrying allele vs the c.1116T>C carrying allele.

We then focused on Family 3 where both c.1327G>A and synonymous c.1080C>T segregate. Blood samples could be collected from the parents (M3, F3) and affected son P18 and daughter P19. As represented in Figure 2B, both children are compound heterozygotes with the maternally inherited c.1327G>A/p.(Val443Ile) and paternally inherited c.1080C>T/p.(Ser360=) that increases Ex10 skipping on minigene assay. As shown, the Ex10^del^ product is over-represented in the 2 patients P18, P19 (both carrying the c.1080C>T variant) compared to the mother (M3) for whom the two bands were of equivalent intensity like in Family 2. Supporting causality of the c.1080C>T in exon 10 exclusion, Ex10^del^ was also found over-represented in the father (F3) sample. This shows that the imbalance of exon 10 skipping induced by variant c.1080C>T is directly detectable from blood RNA samples eliminating the need for skin biopsies.

M3 and both children carry the most common pathogenic variant c.1327G>A which was suspected to cause significant imbalance of the transcripts in favor of the c.1327G allele (Figure 1A, P29). Family 3 was therefore tested for detection of RT-PCR products corresponding to exon 13 (*OCA2*-Ex13) carrying nucleotide c.1327. We successfully amplified a PCR product of the expected size for all members of the family. The nucleotide change c.1327G>A removes a MaeIII restriction site normally present in *OCA2*-Ex13 (Lee et al., 1994). If both c.1327G and c.1327A alleles are represented in a balanced fashion at the blood cell mRNA level, digestion of the RT-PCR products in samples from c.1327(G/A) heterozygotes (M3, P18, P19) should result in both undigested (298 bp) and digested (126+172 bp) products. As shown in Figure 2C, no undigested RT-PCR product could be detected, indicating that too little if any transcripts with variant c.1327A were recovered from the blood cell samples. This was confirmed upon sequencing which identified only wild-type (c.1327G) RT-PCR products (data not shown) from M3, P18 and P19 like from F3 who is homozygous for c.1327G. This suggests that c.1327G>A (allelic frequency 0.003 in gnomAD; most frequent single nucleotide variant in our cohort present in 90 patients out of 414 OCA2 patients) unexpectedly results in a lack of detectable blood cell *OCA2* transcripts.

All in all, we can conclude that the pathogenic imbalance of functional *OCA2*-Ex10^+^ vs. *OCA2*-Ex10^del^ due to identified or yet unidentified variants can be detected directly from blood RNA samples of patients and/or their heterozygous parents. Moreover, these samples can be used to assess the relative representativeness and sequence of transcripts carrying variants located anywhere in the gene.

## DISCUSSION

As much as 50% of disease-causing variants are thought to be pathogenic through splicing anomalies (Baralle & Buratti, 2017), with most of the splicing variants lying outside the canonic dinucleotide splice junction sequences. Because our knowledge of splicing regulation is still limited due to the high complexity of underlying processes, prediction of splicing variants from primary gene sequence is generally poorly reliable. Conclusive genetic diagnosis therefore relies on mRNA analysis, either using a minigene assay, or directly from patient tissue. Minigene assays, although performed in an *in vitro* heterologous cell system, have proven highly reliable to classify suspected splicing variants. However, they require each variant to be produced by cloning and/or site-directed mutagenesis. Moreover, they generally allow to test mis-splicing of only one exon at a time. RNA diagnosis from patient cells or tissue have become widely standardized in recent years (Bournazos et al., 2022). Providing the causal gene is expressed in tissue that is compatible with minimally or non-invasive sampling, RT-PCR products encompassing the suspected mis-spliced exon(s) can be easily obtained, visualized by electrophoresis, and sequenced.

Our large cohort of patients with clinically diagnosed albinism (Lasseaux et al., 2018) provides the unique opportunity to select and test a wide panel of individuals and families for whom genetic diagnosis is pending due to variants of unknown significance or missing variants. Genes implicated in albinism have a more or less specific pattern of expression depending on their role in the pigmentation pathway. For instance, *TYR* (OCA1)*, TYRP1* (OCA3) and *DCT* (OCA8) that encode the 3 melanogenic enzymes are hardly expressed in cells others than pigmentary cells, with no evidence for detectable transcripts in clinically accessible blood samples (GTEx Portal: https://gtexportal.org/ and our own data, not shown). By contrast, transcripts for *OCA2* (OCA2; second most frequent type of albinism) are identified at low levels in whole blood samples indicating that RNA diagnosis should be valuable when genetic diagnosis is inconclusive for suspected OCA2 patients (for instance only one identified pathogenic variant).

As a proof-of-concept of the usefulness of RNA diagnosis for these patients, we focused on a subgroup of OCA2 families with specific emphasis on variants that may induce pathogenic skipping of the in-frame exon 10. *OCA2* exon 10 is highly sensitive to skipping: the wild-type gene is transcribed into a major full-length functional mRNA (Ex10^+^), but a minor form with exon10 skipping (Ex10^del^) is detected in most tissues (Jaworek et al., 2012; GTEx Portal: https://gtexportal.org/). No known function for Ex10^del^ has been reported in the literature, suggesting that, like *CFTR*, *OCA2* has probably evolved under the dual selection of optimized amino acid composition and splicing efficacy (Pagani et al., 2003, 2005; Raponi et al., 2007). Of note, no Ex10^del^ transcript is reported in well-studied pigmented mammals such as rat and mouse (https://www.ensembl.org/; https://www.ncbi.nlm.nih.gov), reinforcing the hypothesis that in Human, Ex10^del^ transcript is a nonfunctional byproduct of evolution.

Among the 6 VUS that were selected for this study, 3 are located in one of the intronic sequences adjacent to exon 10 (at position −10; −9, +6 respectively) which makes them likely to induce mis-splicing. Although only c.1116+6T>C is predicted to affect splicing *in-silico* (Table 2), minigene assay allowed us to functionally illustrate full skipping of exon 10 for all 3 variants (c.1045-10T>G; c.1045-9T<G; c.1116+6T>C) which can therefore be reclassified as pathogenic.

Two of the studied VUS are synonymous. Synonymous variants have long been assumed to be likely benign, i.e. with no consequence on gene expression. High-throughput sequencing has however revealed that synonymous variants can have an effect through different ways, the most common being splicing interference (Anna & Monika, 2018). In most cases, these variants change the first or last nucleotide of the skipped exon adjacent to the consensus splice acceptor or donor site. Such variants considered pathogenic are reported in several albinism genes including *OCA2*. For instance, synonymous substitutions of the last nucleotide of *OCA2* exon 17, 19 and 20 are reported pathogenic relying on robust *in-silico* prediction and/or minigene assay (Lasseaux et al., 2018; Mauri et al., 2017; Rimoldi et al., 2014). The first synonymous VUS in exon 10 tested in this study, c.1047C>T/p.(Ile349=), has been identified in only one patient of our cohort (P6) in *trans* of a pathogenic missense. It has been reported twice as benign in ClinVar. The substitution lies 3 nucleotides away from the 5’ splice site of exon 10, and is not predicted to result in exon skipping by the common *in-silico* tools (Table 2). Still, using the minigene system, we were able to detect a significant increase in exon skipping induced by this variant. Same minigene transcript profile was obtained with the second synonymous variant of exon 10 that we tested, i.e. c.1080C>T/p.(Ser360=). This VUS lies in the middle of exon 10. It was first reported as a polymorphism considered as non-pathogenic by Oetting (Oetting et al., 2005) and is not predicted pathogenic by *in-silico* analysis. As these 2 synonymous variants resulted in consistent but incomplete skipping in the minigene system, we further explored the mRNA profile from patient specimens and confirmed the predominance of non-functional *OCA2*-Ex10^del^ mRNA from both skin biopsies and blood RNA samples.

The last VUS that we tested by minigene assay, i.e. c.1095_1103del/p.(Ala366_368del) is an in-frame deletion predicted to result in a 3 amino acid deletion in the 3^rd^ transmembrane domain of the P protein. The enhanced exon skipping revealed by the minigene assay indicates that the pathogenicity of this variant is related to mis-splicing rather-than or in-addition to the predicted protein alteration.

Importantly, the functional data supported in this paper allows reclassification of 6 variants for which prediction by standard tools (SpliceAI, SPiP) fails to support pathogenicity.

At least two additional synonymous variants in exon 10 of *OCA2* were found in our cohort of patients with albinism but were not included in this study. Variant c.1113C>T/p.(Gly371=) is located 3 nucleotides away from the donor splice site and could therefore be suspected to interfere with splicing of intron 10. This variant is however frequent in healthy population database (2% in gnomAD with 177 homozygotes) and was also found, albeit non-causal, in 8 independent patients of our cohort with a confirmed molecular diagnosis of albinism relying on pathogenic variants in *OCA2* or in another albinism gene. Therefore, this variant can be classified as benign. Variant c.1065G>A/p.(Ala355=) is most common in Europeans with light pigmentation. Genome-wide association studies have suggested that c.1065G>A may increase exon 10 skipping by 20% per allele compared to the reference allele (Crawford et al., 2017). This variant has a high frequency (63% in gnomAD) and has been found homozygous in numerous healthy relatives of our cohort bearing also a heterozygous pathogenic variant of *OCA2* (data available on request). Moreover results from minigene assays by *i.* Jaworek et al. (Jaworek et al., 2012) using the reference allele c.1065G and *ii.* ourselves using the c.1065A allele do not suggest a substantial additive effect of this variant in *cis* to another splicing variant such as the ones we identify in this paper.

This study, designed to assess the usefulness of blood RNA samples analysis for suspected OCA2 patients, primarily focused on splice variants in and around exon 10 of which inclusion, as we show, is highly sensitive to sequence changes. In parallel, the interpretation of RNA profiles obtained from samples of 2 independent cases (P29 with the intronic c.1116+6T>C and P18/P19 siblings with the synonymous c.1080C>T) drew our attention to the effect of the second variant which in both cases was clearly identified as c.1327G>A/p.(Val443Ile). The latter is the most frequent pathogenic variant of *OCA2* (Lewis, 1993) and has been reported in many previous publications (Gargiulo et al., 2011; Hutton & Spritz, 2008). It is also the most frequent pathogenic SNP of our cohort (90 carriers out of 414 OCA2 patients with 11 homozygotes). Interestingly, seven c.1327G>A homozygotes are scored in the gnomAD healthy population database for whom a mild phenotype may have gone unnoticed. Two functional studies have been reported on this variant based on the expression of the mutated cDNA (Sviderskaya et al., 1997; Bellono et al., 2014). They both indicated that the Val443Ile substitution decreases but does not fully abrogate the P protein function. Here, we unexpectedly failed to detect *OCA2* transcripts carrying c.1327G>A, in blood RNA samples from P29, P18/P19 siblings and their parents, all heterozygous for this variant. Whether this missense variant exerts its pathogenic effect in pigment cells by impairing production/stability of *OCA2* mRNAs rather-than-by or in-addition-to amino acid change as previously suggested remains to be determined.

To conclude, we describe a non-invasive RNA diagnostic assay for one of the major OCA gene whose expression, unlike that of *TYR* (OCA1), can be detected elsewhere than in pigment cells. This makes it possible to routinely test many other variants, not only for *OCA2* but also for *SLC45A2*, for which specific transcripts are also detected in blood (data not shown).

Our study allows us to reconsider several VUS of *OCA2* as pathogenic by mis-splicing. We show that this anomaly as well as other types of transcripts imbalance can be detected directly from blood RNA samples, avoiding invasive biopsies. We therefore recommend systematic collection of a blood RNA sample from patients with inconclusive genetic diagnosis and suspected OCA2.

## Data Availability

All data produced in the present work are contained in the manuscript

## ACKNOWLEDGMENTS

The authors are grateful to the French Albinism Association Genespoir for financial support and timeless action in favor of patients with albinism. Patients are warmly acknowledged for their participation in this study. We would like to thank Dr. Patricia Fergelot for providing the pSPL3B plasmid and Dr. Marie-Laure Vuillaume-Winter for the minigene assay protocol. We thank all the clinicians who contributed by addressing patients for diagnosis : Prs. Robert Aquaron, Sophie Blesson, Emmanuel Bourrat, Rodolphe Dard, Sabine Derrien, Valérie Drouin-Garraud, Josseline Kaplan, Didier Lacombe, Valérie Layet, Isabelle Meunier, Sylvie Odent, Ghislaine Plessis, Julia Roux, Xavier Zanlonghi.

---

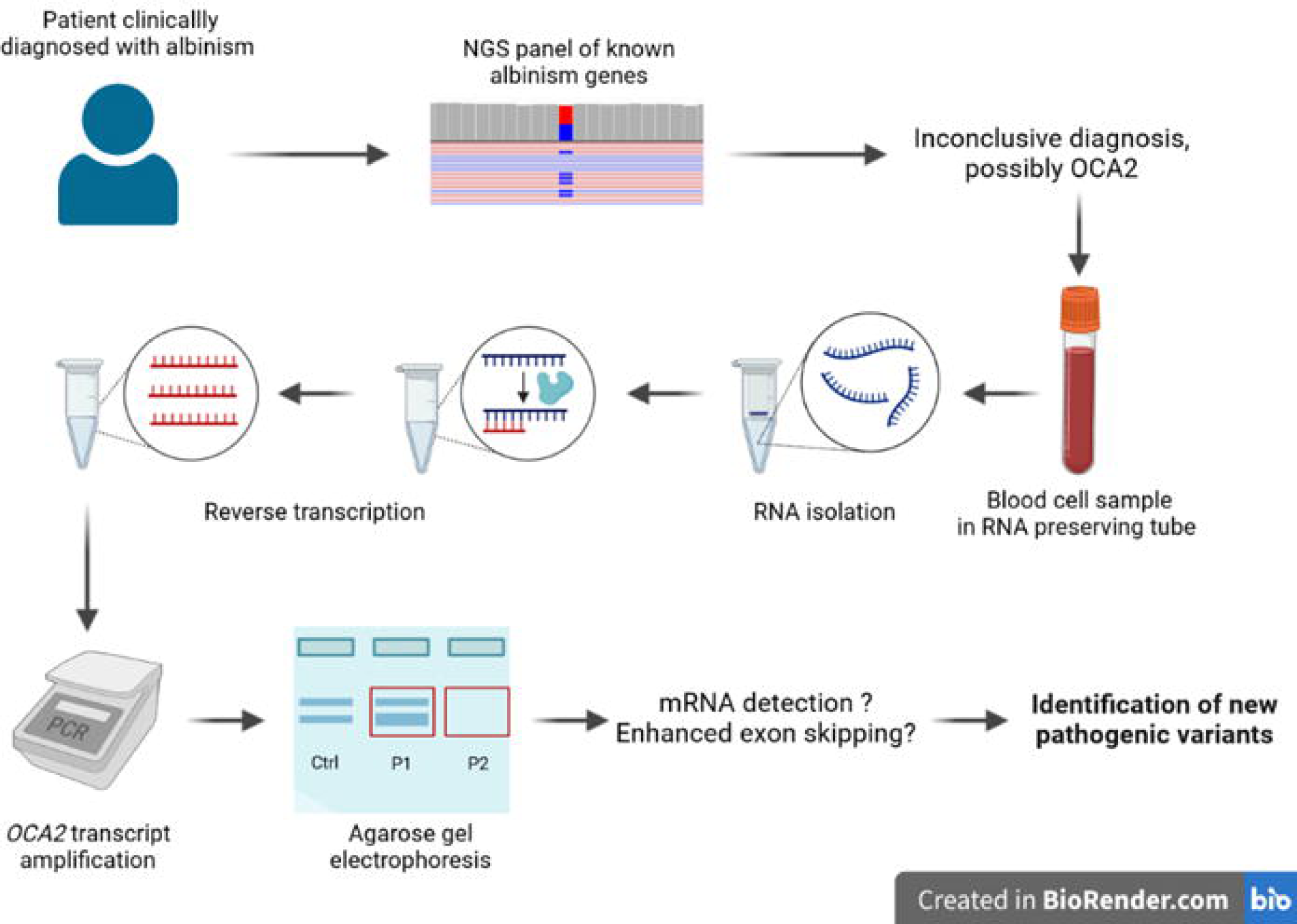

