## Supplemental Table 1 for "Unsuspected consequences of synonymous and missense variants in *OCA2* can be detected in blood cell RNA samples of patients with albinism"

| **Minigene assay** | Homologous recombination in pSPL3B vector | | Forward : 5’ ACCAGAATTCTGGAGCTCGAGCAGTTGCCAAGGGAGCATA 3’ |
| --- | --- | --- | --- |
|  |  |  | Reverse : 5’ ATCCTGCAGCGGCCGCTCGAGCCAAACAGGACACCCTCATC 3’ |
|  | Mutagenesis | c.1045-15T>G | Forward : 5’ GGAACGCGGTAATTTCCGGTGCTTCTTTCCAGATC 3’ |
|  |  |  | Reverse : 5’ GATCTGGAAAGAAGCACCGGAAATTACCGCGTTCC 3’ |
|  |  | c.1045-10T>G | Forward : 5’ GCGGTAATTTCCTGTGCGTCTTTCCAGATCGTGC 3’ |
|  |  |  | Reverse : 5’ GCACGATCTGGAAAGACGCACAGGAAATTACCGC 3’ |
|  |  | c.1045-9T>G | Forward : 5’ GGTAATTTCCTGTGCTGCTTTCCAGATCGTGCAC 3’ |
|  |  |  | Reverse : 5’ GTGCACGATCTGGAAAGCAGCACAGGAAATTACC 3’ |
|  |  | c.1047C>T | Forward : 5’ TGTGCTTCTTTCCAGATTGTGCACAGAACTCTGGC 3’ |
|  |  |  | Reverse : 5’ GCCAGAGTTCTGTGCACAATCTGGAAAGAAGCACA 3’ |
|  |  | c.1064C>A | Forward : 5’ TGCACAGAACTCTGGAGGCCATGCTGGGTTC 3’ |
|  |  |  | Reverse : 5’ GAACCCAGCATGGCCTCCAGAGTTCTGTGCA 3’ |
|  |  | c.1076G>A | Forward : 5’ TGGCGGCCATGCTGGATTCCCTTGCAGCACT 3’ |
|  |  |  | Reverse : 5’ AGTGCTGCAAGGGAATCCAGCATGGCCGCCA 3’ |
|  |  | c.1095_1103del | Forward : 5’ TTCCCTTGCAGCACTGGCTGTGATTGGCGATGTAAGTT 3’ |
|  |  |  | Reverse : 5’ AACTTACATCGCCAATCACAGCCAGTGCTGCAAGGGAA 3’ |
|  |  | c.1103C>T | Forward : 5’ CACTGGCAGCACTGGTTGTGATTGGCGATGT 3’ |
|  |  |  | Reverse : 5’ ACATCGCCAATCACAACCAGTGCTGCCAGTG 3’ |
|  |  | c.1116+6T>C | Forward : 5’ GTGATTGGCGATGTAAGCTGTCACAGTCCCAATCC 3’ |
|  |  |  | Reverse : 5’ GGATTGGGACTGTGACAGCTTACATCGCCAATCAC 3’ |
|  | pSPL3B specific primers for splicing assessment | | SD6: 5′ TCTGAGTCACCTGGACAACC 3′ |
|  |  |  | SA2: 5′ ATCTCAGTGGTATTTGTGAGC 3′ |

| **RT-PCR products cloning from family 1 biopsies** | Forward: 5′ GCAGGTCACTCACAACTGGA 3′ |
| --- | --- |
|  | Reverse: 5′ GACGTGAATCTCGTGCTTCAGTT 3′ |
| **RT-PCR for exon 10 targeting in blood sample** | Forward: 5′ CTCCGCGGAAGTGTAGAAAC 3′ (exon 9) |
|  | Reverse: 5′ GTGAAGAGGAGCATGGTGGT 3′ (exon 13) |
| **RT-PCR for exon 13 targeting in blood sample** | Forward: 5′ TTCAGAAACGGGATTTTTCG 3′ (exon 12) |
|  | Reverse: 5′ CTTCCTCAGCTCTTGGTTGG 3′ (exon 14) |

**Table S1.** Listing of primers.
